# The hidden variable in the dynamics of transmission of COVID-19: A Henon map approach

**DOI:** 10.1101/2020.06.02.20119859

**Authors:** Akshay Pal, Jayanta K Bhattacharjee

## Abstract

We consider the transmission dynamics of COVID-19 which is characterized by two distinct features. One is the existence of asymptomatic carriers which is the hidden variable in the problem. The other is the issue of latency which means that among the symptomatic carriers there could be a fraction whose symptoms develop after a couple of days. We write down a Henon-like map to take these effects into account and find that the map has an unusual fixed point which corresponds to a very high ratio of asymptomatic to symptomatic infected persons. This fixed point has its own stability zone and influences the dynamics overall. Although a rather elementary model, we show that it is not totally devoid of reality.

## I INTRODUCTION

In December 2019 a new epidemic started spreading across the world with two unique features. To recall the simplest scenario for an epidemic, consider a community with two kinds of people (population in fractions) on a given (let’s say the nth) day-*x_n_*, the number of healthy (virus free)but susceptible people and *y_n_* the number of infected people. If we ignore the number of deaths and assume that on a given day the number of new infected people will be jointly proportional to the existing number of infected and uninfected people, we get the logistic map [1] describing the spreading of a standard influenza virus. More sophisticated versions of this are the Ross model [2] and the SIR model [3]. With this new virus there are two dangerous twists.

A. There are two kinds of infected people- one group which does not show any symptoms but can pass it on over the next couple of weeks (i.e. several days). These are the asymptomatic spreaders - the hidden variable in this problem.
B. The other twist is that those who are symptomatic may take a certain number of days (maximum of three or four) to develop the symptoms. Hence this group will have been infecting people not just on the n-th day but also on n-1, n-2 th day if the latency period is taken to be 3 days. This latency period implies that any map (or differential equation) will have to have an in-built delay. The standard literature [4,5] of the dynamics of the spreading of infection has been very nicely summarised in Ref 6. The dynamics is traditionally written in terms of differential equations. The simplest is the logistic equation corresponding to the map described above. The more complicated variety (the analogue of the Lotka-Volterra population dynamics) is the Ross model for spreading of malaria and the popular SIR (susceptible- infected- recovered) developed by Kermach and McKendrick (3). None of these models include the two vital issues mentioned above. A variant of the SIR model which is the stochastic SIR model [7] is an important variation since it includes the process of contact tracing and was used in the early stages of the present diseae. Variations of the stochastic SIR model have been recently studied [8,9]. They too do not explicitly discuss the asymptomatic infected class. More recently a model consisting of a set of delay differential equations was written down which includes all the new features mentioned above [10] Simplified versions of the complete model which are particularly suitable for taking into account the interventions on the part of the government (e.g. imposition of lockdown, requirement of social distancing etc) and analyzing the routes to the decay of the epidemic have been studied in [11]. An interesting variant, starting from a different premise, is the work of Cherednik [12] and a discussion of the early mathematical models pertaining to COVID-19 can be found in Ref [13]. In this work, we will write down a simple Henon like map for the spreading of this infection with the existence of asymptomatic people [14] taken into account but leave out the effect of contact tracing and delay to start with. Our goal is simple. We want to point out that the inclusion of the asymptomatic infected drastically alters the fixed-point structure and very clearly indicates the role of this hidden variable in this problem. We will introduce an extension which takes into account the contact tracing and also explore the existence of the delay in the appearance of the symptoms of the infection for the symptomatic infected. However, all kinds of governmental interventions will be generally left out except when we demonstrate that in confronting reality this simple map is not entirely useless.

## 2. Basic model of Henon map

We take three kinds of people at large on the n-th day–the healthy *x_n_*, the asymptomatic infected *y_n_* and the symptomatic infected *z_n_*. Except for the very early stages there will be a fourth - the number of people who are quarantined- this can be people who are hospitalized or people who are self-quarantined on suspected infection. This is again over –simplified. What we really want to know is the total number of people who have tested positive for the virus on a given day. This again involves including the inherent delay in the testing procedure. For a complete description of the variables involved and the various delays one should consult Ref [10]. We proceed to denote this “removed from scene” variety by *q_n_*. The deaths will be ignored (the percentage generally is small) and we will keep *x_n_ + y_n_ + z_n_ + q_n_* = 1 (constant). There is a catch here. The number *x_n_* denotes the total susceptible. However, a small fraction of x will be the recovered set who presumably will be immune. This fine point we will ignore as nothing about immunity from this virus is known at present. The type of the different population described above will be called X, Y, Z and Q.

X interacts with Y and *Z*. This interaction produces Y with a probability ‘p’ and Z with a probability ‘1-p’. The strength of X and Y interaction is ‘a’, X and Z is ‘b’. On the n+1th day

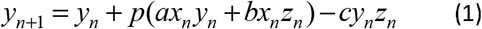

The last term is the Y and Z interaction which send people to class Z with interaction strength ‘c’. There can be another depletion in the class Y. This is through contact tracing. This will be a small effect which we will address a little later. There will be a fraction of Z who will go into quarantine and this fraction is denoted by ‘f and hence

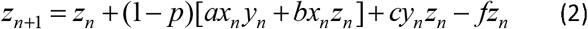

Finally, we have the quarantined set Q who acquire people from the set Z and through treatment or self-recovery restore people to the healthy set X. Consequently,

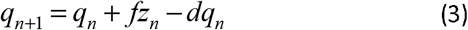

In principle all *z_n_* should be sent to quarantine making *f* = 1. In reality, this may not be so. We will work with a local equilibrium in Q (simplifying assumption to obtain a reasonable two-dimensional map), so that 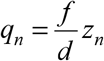. This makes our conservation law constraint give 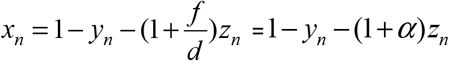 and sets up a two dimensional map. We define b= *ηa, c = aβ* and *f = ag* so that “a” can be an adjustable scale parameter- very useful to make the map appear relevant. Finally, we have our version of the Henon map as

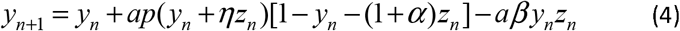

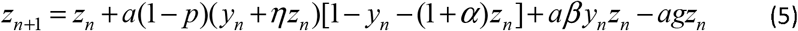

There are two trivial fixed points: *y*_0_ = *z*_0_ = 0 and *y*_0_ = 1, *z*_0_ = 0. The existence of the latter shows the importance of the existence of the asymptomatic cases.

To find the non-trivial fixed point (there are two, only one is positive), we see immediately from Eqs (4) and (5) that the fixed point for y is

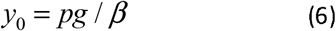

The corresponding *z*_0_ is found from Eq. (4) as the roots of the quadratic

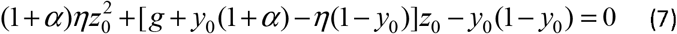

We need to discuss the stability of the fixed points. First linearizing around (0,0)

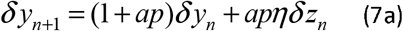

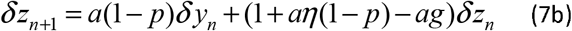

For instability at least one eigenvalue of the stability matrix must have magnitude greater than unity. The eigenvalues of the stability matrix come from the roots of the quadratic equation

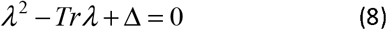

Tr is the trace of the stability matrix and **Δ** is the determinant. For instability we need *Tr >* 1 + Δ The trace is

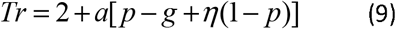

while the determinant works out to be

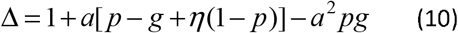

So, the origin is destabilized for *a*^2^ *pg >* 0 which is always true.

Linearizing around the fixed point (1,0).

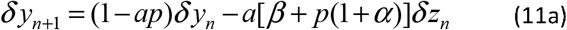

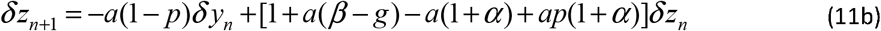

The requirement for the stability of this fixed point, as calculated by a procedure identical to above, is *pg > β*. The fact that this fixed point has a stability zone (see Fig 1) shows its importance in the problem and is really the primary feature of this work. The stability zone makes good sense as it requires the probability of producing asymptomatics in an interaction to be greater than a critical value. It is also favoured if the interaction producing the symptomatic from asymptomatics is small. What this fixed point physically signifies is that the number of asymptomatic carriers is greater than the symptomatic ones by order of magnitude if this fixed point is stable. One can argue that we did not get a finite ratio, we have got something infinitely large and hence is unphysical. As soon as the effect of contact tracing is included this infinite ratio becomes large but finite. Even when this fixed point is unstable it can fashion the course of the dynamics.

**Fig 1:**
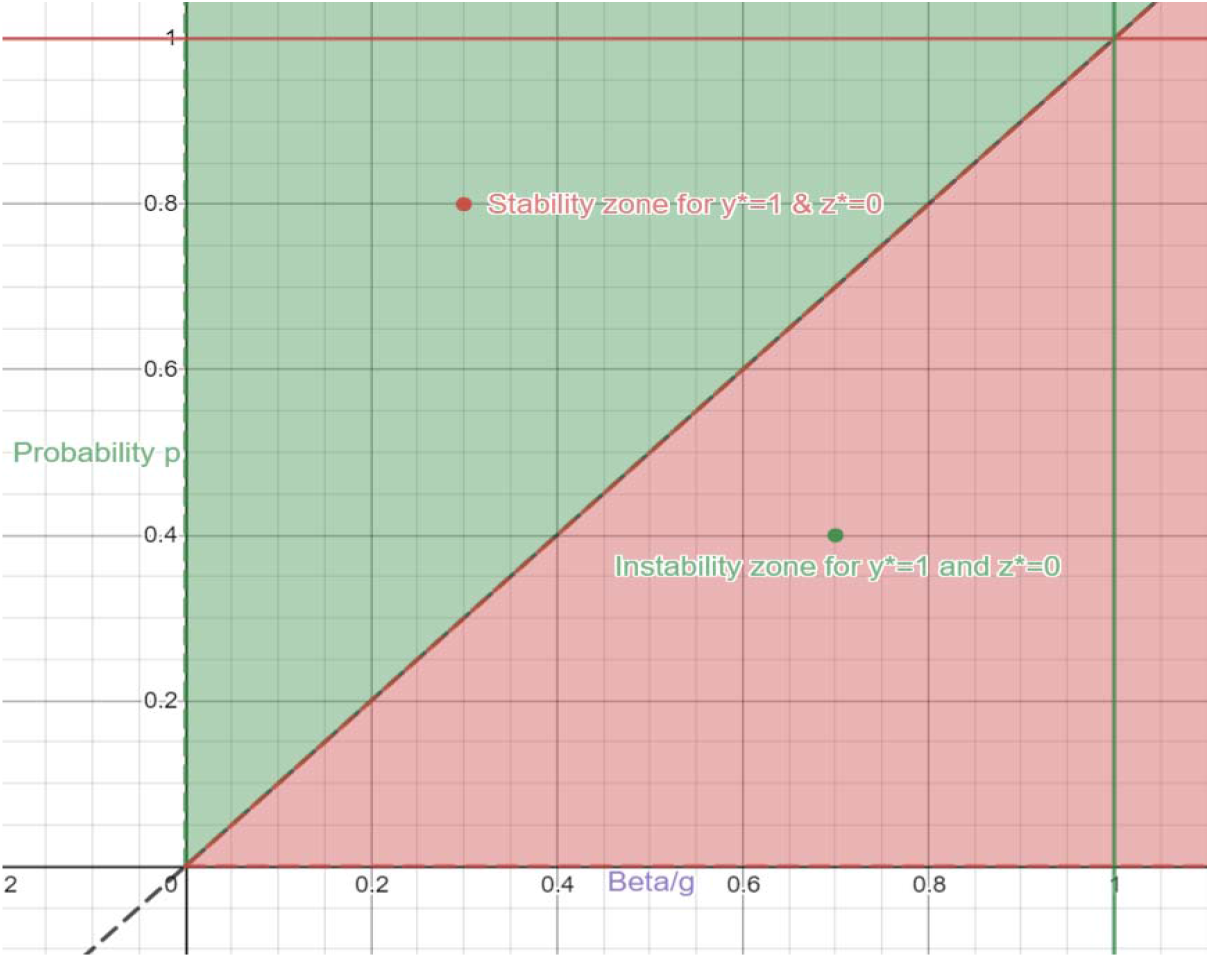
The stability and instability zones of fixed point (1,0)

The important parameters, from Eq.(6) ought to be p,g and a. Moreover, the parameter ‘a’ control sall the other interaction parameters. But actually parameter ‘a’ was the interaction term between susceptible and asymptomatic patients. This shows the importance to include asymptomatics for coronavirus dynamics. We will keep the parameters *η* and *α* fixed at unity. We will consider two cases:

i. *y*_0_ and *z*_0_ comparable.
ii. *y*_0_ significantly greater than *z*_0_.

In Case1, we choose parameters p=0.5, g=0.5 *, β =* **1** and a=0.5 which leads to *y*_0_ = 0.25 from Eq.(6) and *z*_0_ = 0.25 from Eq. (7). We show *z_n_* vs *n* in Fig 2 as Case 1.

In Case 2, we choose p=0.8, *g =* 0.75, *β =* 1 and a=0.5 which lea ds to *y*_0_ = 0.6 from Eq.(6) and *z*_0_ = 0.132 from Eq.(7). The trajectory *z_n_* is shown as Case 2 in Fig 2.

**Fig 2:**
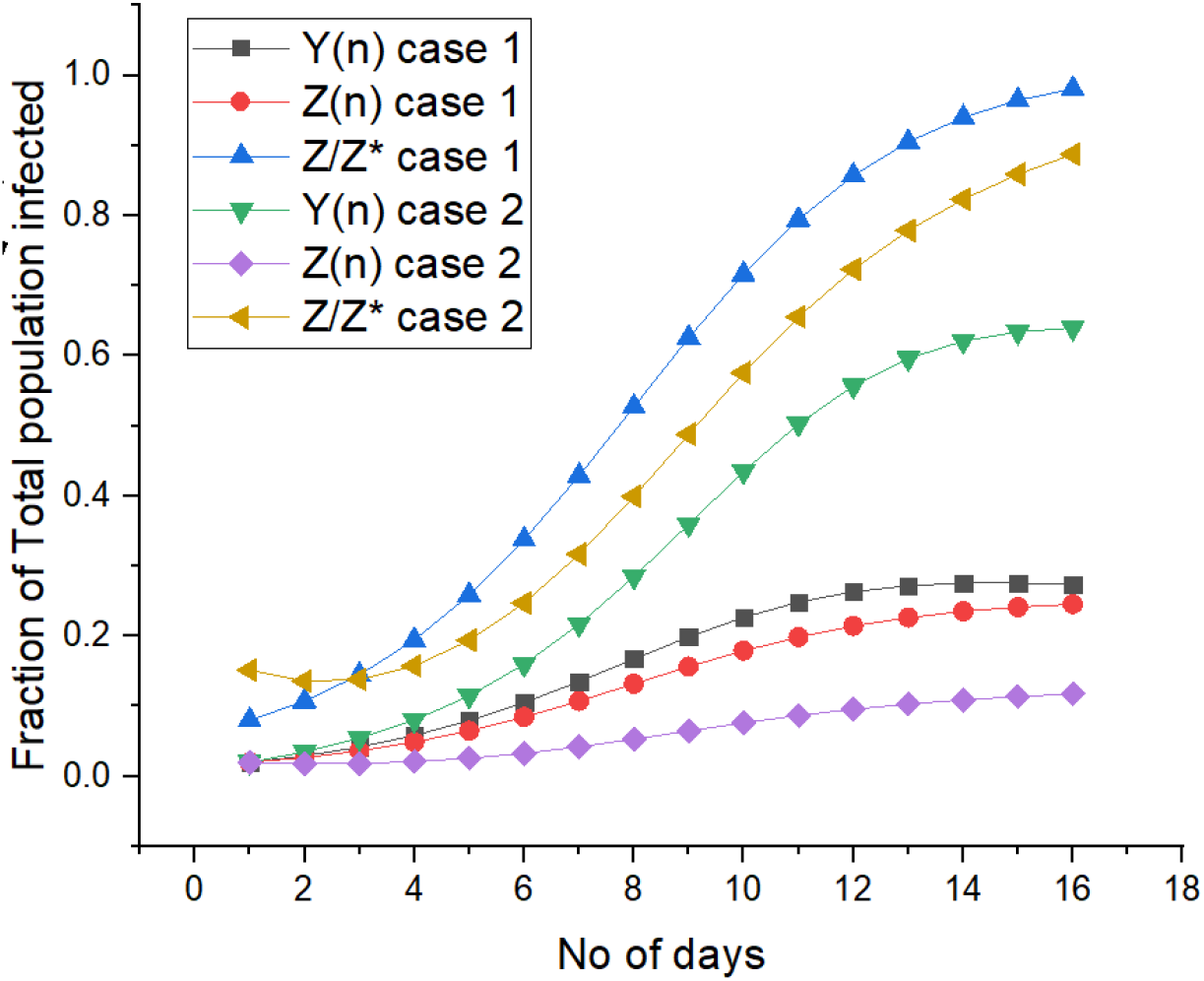
Case 1 and Case 2 are shown above. The parameters for Case 1 p=0.5, g=0.5, and a=0.5. The parameters for Case 2 p=0.8, g=0.75, Where z* is the saturated fixed point for asymptomatics. Z*=0.25 in case 1 and Z*=0.132 in case 2.

The primary difference between the two cases is in the rate at which the saturation is approached. The influence of large number of asymptomatic carriers makes the saturation of the symptomatic infected much slower than in Case 1. This dynamics is likely to show a long-time tail.

We now want to show that the map considered here is not totally irrelevant in practice. To do so, we consider the instance of Austria where an early lockdown was declared by the government and for the most part the growth and saturation of the disease occurred under the same conditions. For a wide variety of choice of parameters similar to the Case1 above, we can get a reasonable description of the time development of the variable ‘z’. This is shown in Fig 3, where we also show the best fit curve to the data.

**Fig 3:**
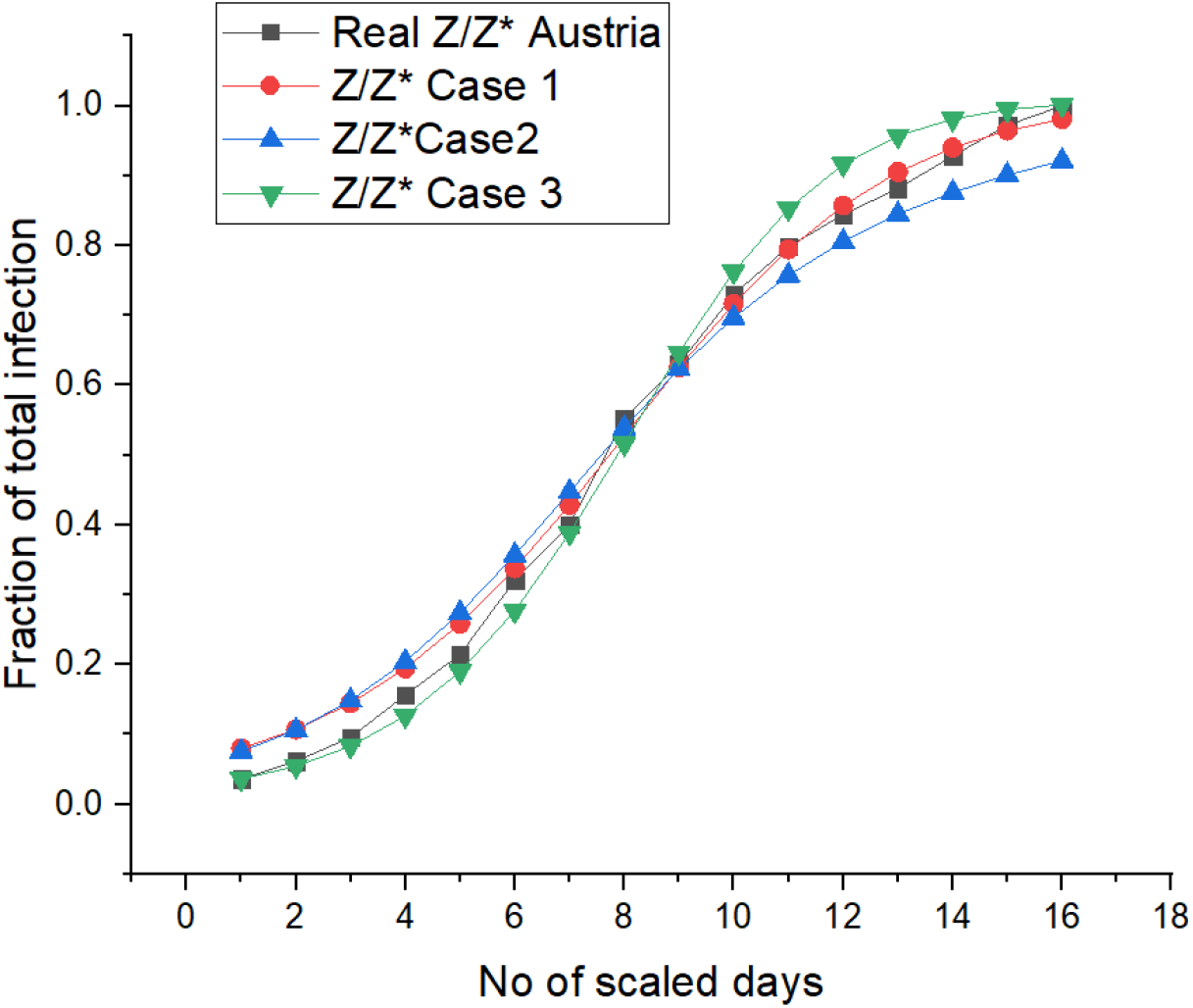
In above graph “Real Z/Z* Austria” represent the actual Z/Z* of Austria from 13 March to 12 April. The days are scaled accordingly. Case 3 is the same as Case 1 only the interaction parameter a=0.65. Before 8^th^ scaled day case 3 overlaps with the actual data of Z/Z* and after 8^th^ day Case 1 overlaps. This clearly shows that the interaction parameter’a’ has reduced from a=0.65 to a=0.5 due to lockdown and governmental policies.

A far more complicated is the situation in South Korea where in the very early stage, the Shincheonji gathering produced an unusually fast exponential decay which sent the new cases up to 5000 very soon (about 20 days). Government intervention then slowed the growth and eventually it saturated at a value slightly above 10000 in the next 40 days. Hence in terms of the normalized *z_n_* / *z*_0_, the half way mark is reached incredibly fast and then the rest proceeds like Austria. Consequently, to understand this data, we need to have an “extreme” set of parameters the first 20 days and then switch to Austria-like parameters in the next 40 days. This is what we show in Fig 4.

**Fig 4:**
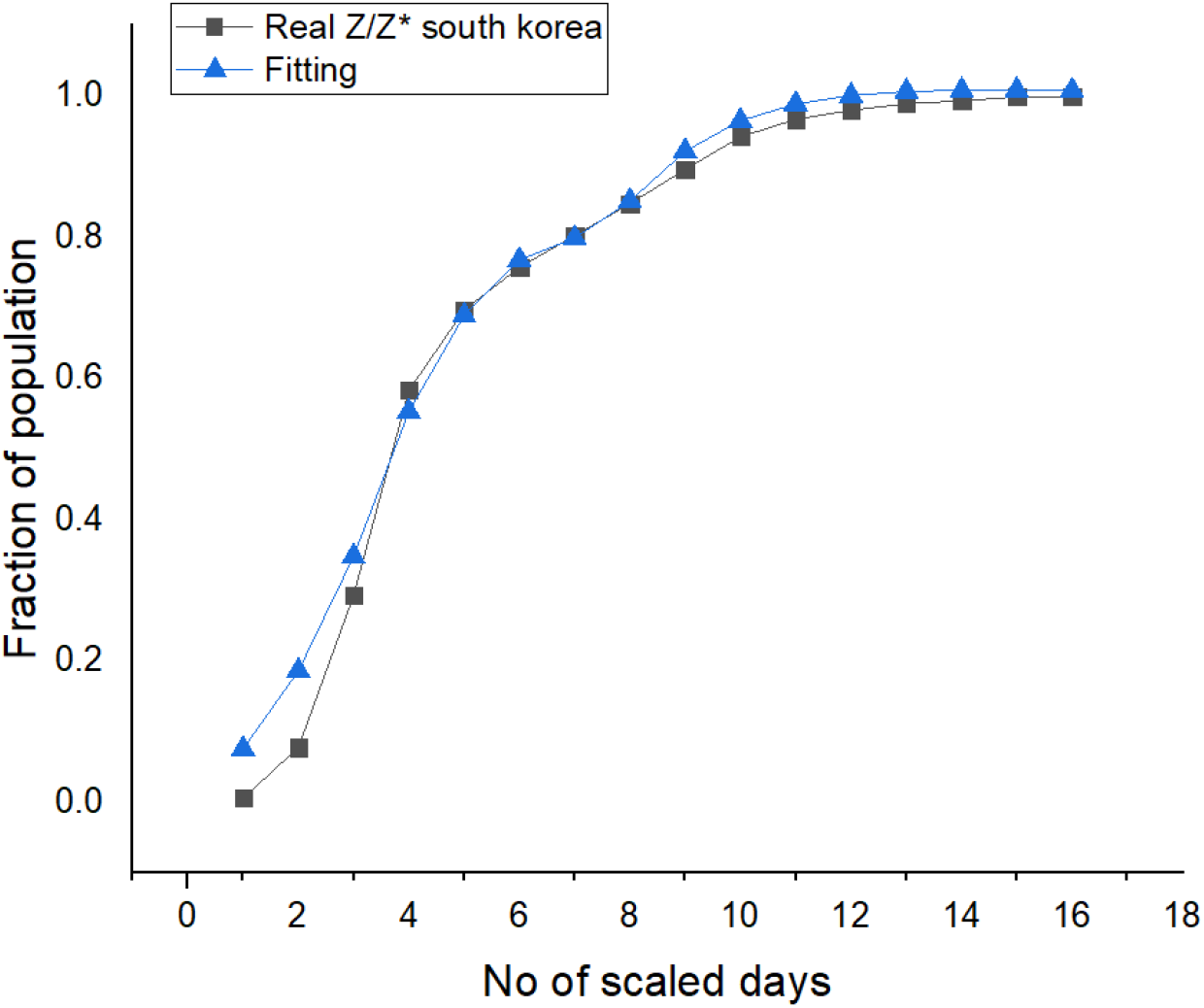
In above graph “Real Z/Z* South Korea” represents the actual value of Z/Z* of South Korea from 12 Feb to 14 April. Days are scaled accordingly. Before 4^th^ scaled day, parameters are difficult to predict due rapid fluctuations of parameters because of the Shincheonji gathering but it can be approximated to p=0.01, a=0.9, g=0.2, On 5^th^ scaled day onwards probability of getting asymptomatics increased p=0.5 because of lockdown and government policies. On 6^th^ scaled day interaction parameter a decrease from ‘a’=0.9 to a=0.8. On 7^th^ scaled day ‘a’=0.7. Finally, on 8^th^ scaled day onwards interaction parameter ‘a’ decreases to a≈0.6 and remains constant afterwards.

## 3. Modification to include contact tracing

The way things work in reality actually changes the fixed point (1,0) to something where the ratio of asymptomatics to symptomatic is large but finite. This is because while members of the set Y are generally not quarantined- an exception is when they are tested due to contact tracing. This implies that in Eq.(4) there will be an additional term which we call *−aμy_n_* and an identical term in Eq.(3) with the opposite sign keeping the conservation law intact. This modification changes Eq.(4) and Eq.(5) to

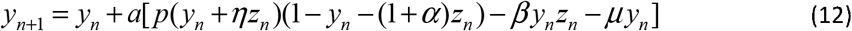

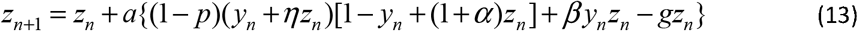

The correction term *μy_n_* is small as the coefficient is small-contact tracing contribution is a second order effect. As is obvious the fixed point (0,0) is unchanged but the fixed point (1,0) will be shifted by a term of *O(μ)* at the lowest order. We write

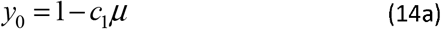

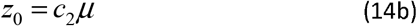

where *c*_12_ are positive coefficients of order unity. From the fixed-point conditions for Eqs (12) and (13), we obtain (we work with *α =* 1 to reduce the number of coefficients)

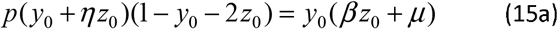

and

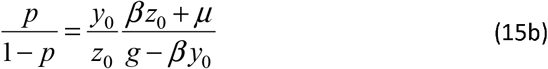

This immediately leads to

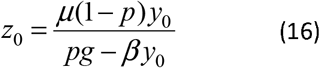

Since the Rhs of Eq.(16) is already *O*(*μ*), we can use just the first term of the Rhs of Eq.(14a) to obtain

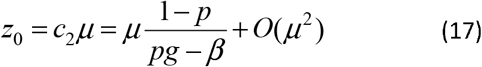

We need *pg > β* for to be right. To find the exact equation satisfied by *y*_0_, we need to insert *z*_0_ as given by Eq. (16) into Eq.(15a) and obtain for the non-zero fixed point values,

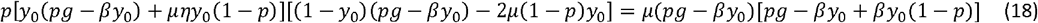

In the case where *μ =* 0, we correctly obtain

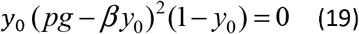

The correction to *y*_0_ = 1 to the lowest order is obtained by inserting the form of Eq. (14a) in Eq. (18) to arrive at

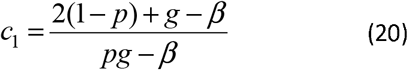

We thus see that including the effect of contact tracing leads to a final state where asymptomatics can be much larger than those with symptoms and this state exists and is stable for the region shown in Fig (1). Hence in reality, the fixed point (1,0) is a limiting case of a more realistic fixed point where the asymptomatic fraction is much greater than the symptomatic fraction with both numbers’ finite. The other positive valued fixed point discussed in Eqs (6) and (7) is also slightly shifted.

## 3. Modification to include Latency

We now discuss briefly the question of latency in the symptomatic carriers. It is possible that among the contributions to *y_n+_*_1_ and *z_n+_*_1_, apart from interactions of the variety *x_n_z_n_* there could be interactions with the z variety of the days n-1, n-2 and probably n-3. These would be interactions of the set X with some fraction of the numbers *z_n−_*_1_, *z_n−_*_2_ and *z_n−_*_3_. These terms which are of the form *x_n_z_n−i_(* i=1,2 and perhaps 3) can be approximated by one representative term *x_n_z_n-1_* with an effective coefficient in a minimal model. An identical argument holds for the generation of *z_n_*_+_**_1_** from interactions between the set Y and the set Z. Consequently, the structure of Eqs (4) and (5) become [we use the parameters *η =* 1*, α =* 1]

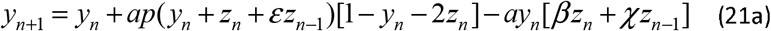

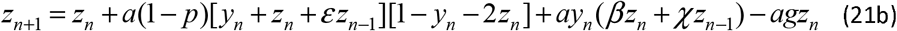

In the above *ε, χ* are two new parameters describing the strength of the latency effect. The fixed points *y*_0_ = *z*_0_ = 0 and *y*_0_ = 1, *z*_0_ = 0 are unchanged while the non-trivial fixed point becomes *y*_0_ *= pg/(β + χ*) with *z*_0_ following from [*y*_0_ + *z*_0_(1 + *ε*)][1 -*y*_0_ -2*z*_0_] = *gz*_0_. The linear stability of the fixed point is now obtained from the roots of a cubic equation. To understand this in the simplest case, we need to look at the (0,0) fixed point. The linear stability equation will now involve a term in *δz_n_*_-1_ in each of Eqs (7a) and (7b) and these terms will be of *O*(*ε*) or *O*(*χ*). This means that in addition to the stability matrix of Eqs.(7a) and (7b), we will have two more matrices one involving the first and third columns and the other the second and third columns. The determinants of these matrices enter the stability analysis and are clearly of *O*(*ε*) or *O*(*χ*). Straightforward algebra leads to a cubic having the form *λ*^3^ *− λ*^2^*Tr + λ*(Δ *+ O(ε, χ) + O(ε, χ*) = 0 The additional root that we get is clearly small and cannot cause instability. Hence, we need to find the first correction to the root greater than unity and that correction is positive ensuring instability of (0,0).

To explore the role of latency on the evolution of *z_z_*, we have solved Eqs(21a) and (21b) corresponding to our Case 1 in Fig2 with three different values of *ε* and *χ*. This is shown in Fig 5.

**Fig 5:**
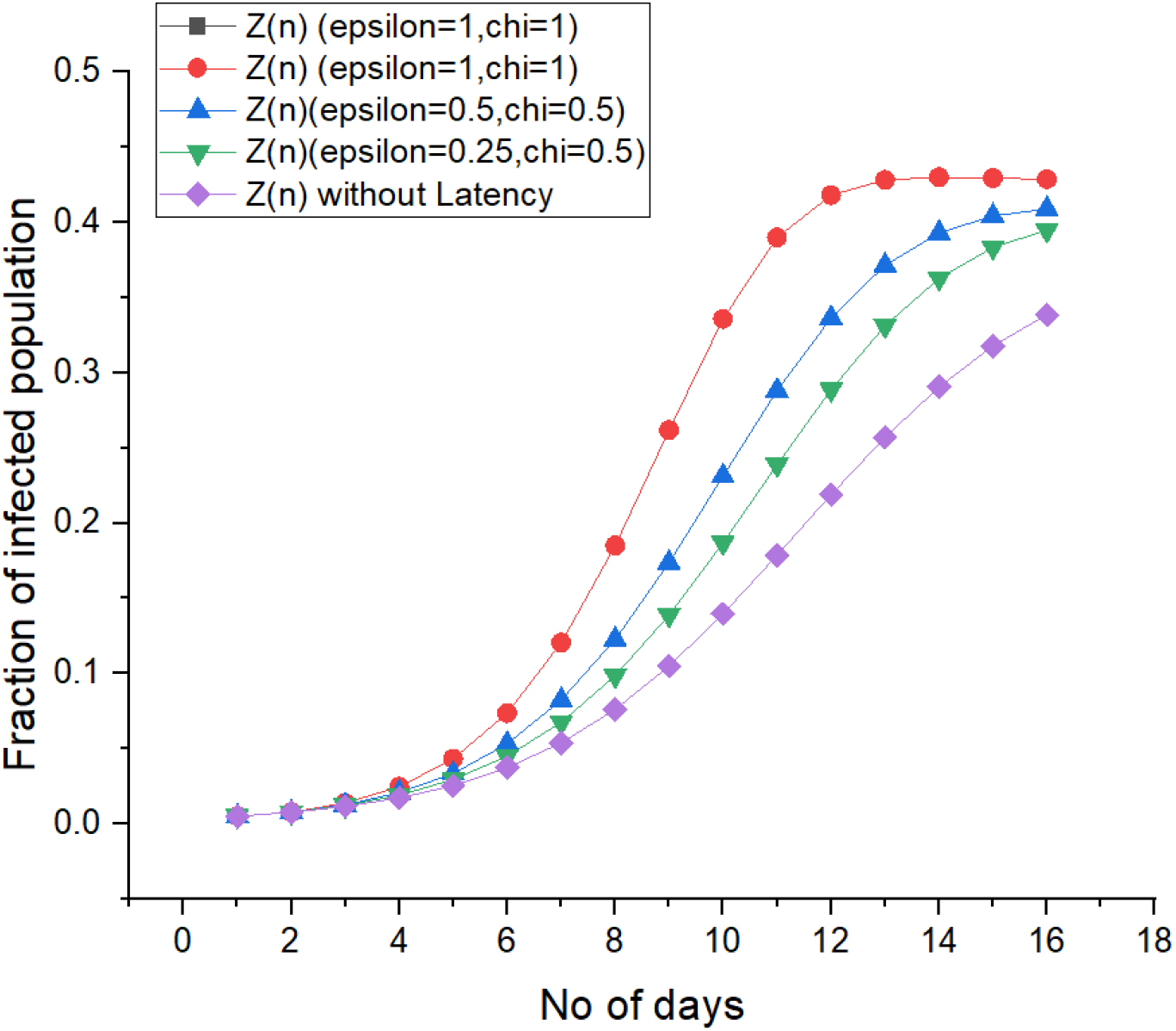
Above graph shows the effect of latency. When the saturation is more rapid, growth rate is high as compared to other two case when and

## 4. Conclusion

We have written down a Henon map for the transmission dynamics of COVID-19 which includes its two basic features- the existence of asymptomatic transmitters and a possible latency period for the symptomatics. The existence of an unexpected fixed point with a large ratio of asymptomatic to symptomatic influences the transmission dynamics strongly. In simple enough situations, the map is capable of producing a reasonable account of the observed dynamics.

## Data Availability

https://www.worldometers.info/coronavirus/

